# Why Primary Care Clinicians use Advice and Guidance: A qualitative study

**DOI:** 10.64898/2026.03.13.26348141

**Authors:** Alice Faux-Nightingale, Rosie Harrison, Claire Burton, Ram Bajpai, Lorna E Clarson, Tina Hadley-Barrows, John Haines, Toby Helliwell, Samantha L Hider, Clare Jinks, Kelvin P Jordan, Natalie Knight, Christian D Mallen, Kayleigh J Mason, Victoria K Welsh

## Abstract

**Background:** Advice and Guidance (A&G) enables primary care clinicians to seek specialist input, supporting decision making and avoiding unnecessary referrals. The use of A&G has significantly expanded, accelerated by COVID19 and contractual changes. While A&G is intended to streamline elective care, concerns persist regarding workload shift, variable responsiveness, and system usability. Despite growing policy emphasis, little is known about why clinicians choose to use A&G.

**Aim:** Explore the current use of A&G within primary care, focusing on decision making processes which underpin PCCs’ decision to use A&G.

**Design and Setting:** Qualitative study set in English Primary Care

**Method:** Twenty semi structured video interviews were conducted with primary care clinicians purposively sampled for maximum variation. Topic guides were developed with PPIE input and refined iteratively. Data were analysed using reflexive thematic analysis within an interpretive description framework, with themes developed collaboratively and refined through discussion with researchers and PPIE contributors. Ethical approval was obtained (REC 333799).

**Analysis:** Four overarching themes encapsulate clinicians’ decisions to use A&G: clinical presentation (acuity and complexity), navigating healthcare pathways, previous experiences of A&G, and using A&G to validate clinical decision making. Barriers included delayed responses and uncertainty about inequitable workload distribution. These factors shape how effectively A&G could be integrated into routine practice.

**Conclusion:** Primary care clinicians use A&G to support patient care and aid decision-making, but its effectiveness depends on timely, clinically helpful responses. Ensuring responses remain appropriate to primary care remit and capacity will be essential if A&G becomes the main route into elective care.

**How this fits in (<4 sentences):**

- Advice and Guidance (A&G), introduced in 2015, enables primary care clinicians to obtain specialist input for patients within elective care pathways without initiating a formal referral; the proposed 2026/27 GP Contract aims to expand or mandate its use in line with the NHS ten-year plan and strategic efforts to move more care from hospitals into the community.
- Our findings suggest clinicians decide to engage with A&G when they can expect timely, high-quality responses that support safe management of sometimes clinically complex patients.
- Primary Care Clinicians are also sensitive to the risks of inappropriate transfer of workload from secondary to primary care, clinical responsibility for conditions traditionally managed in secondary care, and associated costs into primary care, and that these concerns directly influence their willingness to use A&G.
- Ensuring that electronic A&G systems support transparent, two-way communication and maintain visibility for both primary and secondary care teams may therefore improve usability and sustain clinician engagement.

## Introduction

In the UK, primary care clinicians (PCCs) working in the National Health Service (NHS) use Advice and Guidance (A&G) to obtain specialist input to support patient care within elective care pathways. A&G is usually delivered via the electronic Referral System e_RS, a digital interface between primary and secondary care.^1^ The e_RS supports secure, documented communication between primary and secondary care clinicians (SCCs) as electronic messaging with the potential for attachments to support the query.^2^ Telemedicine portals also exist, enabling contemporaneous telephone and electronic discussions with SCCs.

A&G allows primary care clinicians to seek specialist advice to inform shared decision-making and patient management in the community, potentially avoiding the need for an outpatient appointment. It is a key component of the UK Government’s 10-Year Health Plan for England, designed to support outpatient transformation and reduce the number of referrals to secondary care^3^ and shift care from hospital to community.

While there is limited published research evaluating the impact and outcomes of A&G, case based examples are promising.^4^ A&G is purported to support elective care transformation, reducing NHS waiting lists^3, 5^ and referrals^6^; A&G is also reported to support the rapid^6^ timely management of conditions needing specialist^7^ input; shorten the patient healthcare^8, 9^ journey by avoiding unnecessary appointments with associated^10^ reductions in time and financial burdens for patients; and reduce burden on primary care.^6^ There are, however, concerns amongst clinicians in primary care over the potential for workload shift from secondary care^11^ and the impact on PCC autonomy. Although specialty teams work to respond to A&G requests in a reasonable timeframe, increasing volumes of A&G^12, 13^ mean that this is not always possible, leading to delayed responses and associated potential impact on patient care.^14^ Additional concerns have also questioned whether A&G always represents an effective use of clinical time and workload allocation,^14-17^ whether the supporting IT systems are effective,^13^ and whether use of A&G might negatively impact on a patient’s perception of primary care clinicians’ competence.^14^ Though further research is needed to expand understanding of these observations and monitor long-term outcomes.

Use of A&G has increased across specialties,^2, 10^ both due to rapid expansion during COVID-19 pandemic, and the decision of some localities to make the process compulsory^18^ for primary care clinicians making routine referrals. In addition to this, an incentive scheme has been implemented in the UK to encourage primary care clinicians to use A&G, offering £20 for each A&G request submitted from April 2025.^19^ At the time of writing, this incentive is being removed and incorporated into the global sum;^20^ however, the terms of the 2026/27 contract for general practice are currently being contested.^21^

Evidence on the use of A&G, particularly why PCCs choose whether or not to engage with it, remains limited. As uptake expands, understanding how clinicians experience and engage with A&G is increasingly important. Exploring their perspectives can provide insight into how the system operates in everyday practice, including the key facilitators and barriers to effective use. These findings can help inform service improvement and support clinicians as A&G becomes more embedded in primary care.

This study is underpinned by the pragmatist research paradigm, problem centred and practice oriented to bring about change in quality care. We, a group of healthcare researchers and academic clinicians (GPs and Consultant), use reflexive thematic analysis^22^ within an interpretive description^23^ methodology, taking an applied, clinical perspective and acknowledging that we look to develop findings which can be applied to clinical practice, healthcare commissioning and policy development.

The aim of this study, for the first time, was to explore the current use of A&G within primary care, focusing on decision making processes which underpin PCCs’ decision to use A&G.

### Methodology

The study received ethical approval from the North East – Tyne & Wear South Research Ethics Committee (333799).

The COREQ checklist^24^ (Supplementary material 1) and Reflexive Thematic Analysis Reporting Guidelines^25^ have been used to support drafting this manuscript.

#### Recruitment and sampling

Twenty general practices were recruited to the study with the support of two Regional Research Delivery Networks (RRDNs): North West (NW) and West Midlands (WM). Invitations to take part in an interview were distributed to all PCCs working in these practices who used A&G. Interested PCCs returned a reply slip to the study team to aid sampling. Purposive sampling with a focus on maximum variation^26^ was used to recruit a broad range of participants, sampling on demographics age, ethnicity, gender, A&G usage, geographical location, clinical role, and experience in role.

#### Data collection

Data collection took place between October 2024-February 2025 by AFN and RH (PhD) (female, non-clinical qualitative researchers with backgrounds in the social sciences).

Twenty semi-structured interviews were conducted via video-call and recorded using MS Teams. Respondents were not previously known to the researchers. Nineteen interviews were one to one; one GP also asked their practice manager to support them in their interview.

Interview topic guides (Supplementary material 2) were developed with our PPIE and clinical stakeholder groups and were used flexibly, being iteratively revised based on early findings and key policy changes published over the course of data collection. As Reflexive Thematic Analysis is not compatible with concepts of data saturation,^27^ our sample size was determined by the number of practices involved in the study (n=20) and we aimed to interview one PCC from each practice.

PCCs provided consent before the interview, which was reaffirmed during the interview. PCCs were reimbursed at NIHR rates for their time, in the form of a voucher if the interview was conducted outside of worktime or reimbursement to their practice if the interview was conducted during worktime.

Interviews were either professionally transcribed verbatim or transcribed using the in-built transcription function in Microsoft Teams.

#### Data analysis

Interpretive description^23^ is a qualitative methodology widely used in healthcare research to explore experiences and perceptions in clinical contexts. It acknowledges the clinical expertise within the research team and supports an applied, practice-oriented focus, aiming to generate insights that can inform future inquiry and clinical practice. This approach was used to guide the overall design of the study and was complemented by reflexive thematic analysis, which involves systematic familiarisation, coding, and iterative development of themes and subthemes. AFN and RH independently coded transcripts and generated preliminary findings which they shared and refined together to construct research themes. These themes were shared with the wider research team, including academic GPs, who brought additional experience and context to the analysis, and were refined further based on commonality, significance, and relevance to the research questions. Findings were discussed and finalised with the PPIE group and other stakeholders.

#### Patient and public involvement

Members of the public have been involved throughout this study, from the pre-grant development to the dissemination of study findings. The PPIE groups contributed to the development of the research project, contributed to the topic guides and public-facing documents, and supported the research team with the development of the research findings, plans for dissemination, and public-facing dissemination materials.

### Analysis

#### Participant demographics

Thirty nine PCCs expressed interest in participating in an interview (16 from WM, 23 from NW). Twenty participants were sampled and interviews undertaken between October 2024 and February 2025, ten from WM and ten from NW. Interviews took between 29 and 70 minutes, participant characteristics shown in Table 1.

**Table 1:** Participant characteristics.

| ID | Gender | Age (years ) | Job role | Time in role | Frequency of Advice & Guidance use | Practice Indices of Multiple Deprivatio |
| --- | --- | --- | --- | --- | --- | --- |

|  |  |  |  |  |  | n<sup>26</sup> IMD<br>decile<sup>a</sup> |
| --- | --- | --- | --- | --- | --- | --- |
| G01 | F | 30-34 | GP Partner | 5-10<br>years | Weekly | 9 |
| G02 | F | 40-44 | ANP | 0-5 years | Weekly | 9 |
| G03 | M | 45-49 | GP Partner | 20-24<br>years | Very little | 2 |
| G04 | F | 50-54 | GP registrar | 5 years | Weekly | 2 |
| G05 | M | 55-59 | Salaried GP | 30+ years | 3 x week | 1 |
| G06 | M | 50-54 | GP partner | 6-10<br>years | Weekly | 9 |
| G07 | F | 30-34 | GP registrar | 0-5 years | Fortnightly | 3 |
| G08 | F | 35-39 | Salaried GP | 6-10<br>years | Fortnightly | 5 |
| G09 | F | 25-29 | Senior PA | 1-5 years | Fortnightly | 2 |
| G10 | M | 35-39 | Salaried GP | 1-5 years | Daily | 2 |
| G11 | F | 30-34 | Salaried GP | 1-5 years | Weekly | 3 |
| G12 | F | 50-54 | GP partner | 6-10<br>years | Once every 3<br>months | 4 |
| G13 | F | 55-59 | GP partner | 30+ years | 3 x a month | 4 |
| G14 | M | 30-34 | Salaried GP | 0-5 years | 2-3 times a<br>month | 5 |
| G15 | M | 40-44 | Salaried GP | 1-5 years | Weekly | 9 |
| G16 | M | 45-49 | Salaried GP | 11-15<br>years | Monthly | 3 |
| G17 | M | 40-44 | GP partner | 6-10<br>years | Weekly | 6 |
| G18 | F | 45-49 | Locum GP<br>(previously a<br>partner) | 21-25<br>years | Monthly | 4 |
| G19 | M | 45-49 | GP partner | 6-10<br>years | Monthly | 1 |
| G20 | F | 25-29 | Clinical<br>pharmacist | 0-5 years | 2-3 times a<br>month | 3 |

F female, M male, GP General Practitioner, ANP Advanced Nurse Practitioner
<sup>a</sup>Where 1 represents the most deprived decile and 10 represents the least deprived decile

## Analysis

Four themes influenced PCCs’ decisions to use A&G: clinical presentation and the need for specialist input; care navigation and negotiating healthcare systems; experience of A&G; and clinical validation and patient reassurance. Themes are presented below with exemplar quotations.

### 1. Clinical presentation and need for specialist input

Clinicians reported that decisions to use A&G, rather than other referral routes, were predominantly driven by the patient’s clinical presentation and characteristics including age, comorbidity and clinical complexity. Some PCCs felt that complex cases should be seen in specialist clinics whereas others felt that A&G had a role in supporting such cases. Urgency of referral also influenced decisions, with PCCs generally using A&G for non-urgent queries.

> *“…those who are a bit more comorbid or have more complicated issues, I’m more likely to use a bit of advice and guidance for*.*” G14, GP*
>
> *“If it’s that urgent, I’m going to be referring them or sending them up. These are all sort of questions where you’re not quite sure what to do, type things so they’re not an urgent problem” G13, GP*

PCCs used A&G when the clinical scenario was beyond their scope of practice with no reasonably accessible accepted guidance, or where no in-house expertise existed.

> *“We do have primary care guidelines from NICE*…*But there might be situations where seriously there isn’t a guideline for it, so where do you go? And you don’t want to make the wrong decision or fob the patient off* …*So when I’m really worried about patients and I feel worried about sending them home without a clear diagnosis or management or I have zero clue about what needs to be done here, that’s when I really do use A&G*.*” G04, GP*
>
> *“So it’s just the ones in between where you can’t get advice from your partners or colleagues*.*” G13, GP*

### 2. Care navigation and negotiating healthcare systems

A&G was commonly requested to obtain guidance on navigating local healthcare systems, and clinicians described its role in negotiating the interface between primary and secondary care. PCCs reported that A&G helped them identify the most appropriate pathway for initiating specialist care and request interim support for patients awaiting outpatient appointments, while working within system constraints and requirements.

Decisions to initiate A&G were often driven by the need for clarity about local referral pathways, such as which pathway was most suitable for a given patient, given expected referral timeframes, and whether secondary care might convert an A&G query into a formal referral while providing interim advice.

> *“Occasionally what happens is that I’m not really concerned about the clinical element of it, but exactly how do we go about getting things done?” G16, GP*
>
> *“I just feel as a GP I’m forever-I’m forever having to fight the fight the system. No, not fight the system. Navigate the system to get the patient seen in the right place…” G0*3, GP

PCCs commonly used A&G to ask questions pertaining to medicines management, obtain interim management for patients who had already been referred, and for advice confirming the remit of primary care when managing patients in the community.

> *“And I think like majority of mine are like Parkinson patients and specific like neurology where we just do not touch the medications like Cocareldopa*… *As well as that, mental health medication such as like the [name] centre, so about like Olanzapine and Risperidone all these different like you know amber drugs, we just seek advice and guidance*.*” G20, Clinical pharmacist*
>
> *“I had another one this week that was, a man who’s got an overactive thyroid. And I’ve already done a referral, but I wanted to see if he should be starting some medication without having to wait for sort of six weeks*…*” G01, GP*

In some areas, local practices and systems mandated the use of A&G leaving PCCs no choice but to use it:

> *“But I have used it for other things as well. Some of that might be affected by local referral practices and systems that we have in place*.” G12, GP
>
> *“There was a time when the paediatricians mandated that if we’re gonna do bloods for a child*…*We had to ask for advice and guidance. Almost asked for permission before doing those bloods, even if we felt it was clinically indicated*…*” G08, GP*

In other areas, A&G access was limited because services had withdrawn after becoming overwhelmed by demand.

> *“We used to use it for a lot for dermatology lesions, but they’ve now said they don’t have the capacity to manage that*…*” G17, GP*

Regardless of incentivisation, PCCs did voice concerns about the financial implications of care traditionally being delivered in secondary care, being transferred to primary care, for example, the cost of ordering investigations and time and clinical responsibility interpreting and communicating findings.

> *“You see there’s another element to it sometimes because it’s completely not related to clinical knowledge, it’s related to funding and commissioning as well[…]So referral covers not just patient being seen, but also consultants conducting appropriate investigations. So what’s the point of me doing these investigations if I’ve already paid for consultants doing it?” G16*, GP

### 3. Experience of Advice and Guidance

Primary Care Clinician’s decisions to use A&G were shaped by previous experience of using the process, particularly in terms of it working efficiently and finding it clinically helpful. The speed of previous responses from a speciality was a key consideration in using A&G. Where timely responses were routinely received, A&G was perceived as more beneficial for patients than waiting for an outpatient’s appointment:

> *“So I think the ones I’ve used have been very good. You get a very quick response*.[…*] The patients, I mean for gynaecology, I could refer her and she could sit and wait on the 9 to 12 month waiting list to be told what I could have had in 48 hours. So it makes a difference to patient journey as well really*.*” G13, GP*

However, timeliness of A&G responses varied, with most PCCs reporting they did not know an exact timeframe in which to expect a response. Longer response times were reported as detrimental to patient care, introducing delay and contributing to GP workload.

> *“So I can remember once receiving, I’m not going to name the specialty, I got the answer back six weeks later[…]You know for six weeks, the patient’s waiting. We’re waiting. We’re thinking. Should we just refer? […] And it’s also because they’re technically on still on your-, I guess on your workflow because you’ve got them still there as a pending unresolved medical issue, so you have to keep following up with them to check that they’re OK[…]So there are at least one or two extra appointments that they didn’t need, had we just got the advice back and good time*.*” G10, GP*

Experience of long A&G response times (specialty dependant) determined PCC decisions not to use the service again:

> *“[…]Like I did one the other day for [specialty], not heard a thing. It’s not good, is it? So I’m less likely to use it again you see. I’m less likely to use it, you know, next time*.*” G18*, GP
>
> *“…the other day I did one for [specialty] that I’ve never used before and I didn’t hear back for weeks. It took lots of prompting you know and they didn’t. So I probably wouldn’t bother again*.*” G06, GP*

Useful advice from secondary care served as an educational opportunity for PCCs, and there were reports of saving generic advice to use in future, avoiding the need for similar A&G requests:

> *“Most times the advice is pretty is very useful. It’s like educational tool for me[…]I try and save it for me so I can use that for future patients who can present with the same problem*.*” G15, GP*
>
> Though PCCs highlighted examples of A&G use that had benefitted patients, some were hesitant to increase use, describing increase in workload through work shift of tasks traditionally owned by secondary care:
>
> **“***You find it useful with one [specialty] or it’s quite useful to A&G for this thing so I’ll do it for some other speciality and then before you know it[…] you can do more for the patient so I guess that’s sort of changing your role a bit; not your role, but what you feel that you can do. But then when you stop and look around, you think, ‘Wow! Why am I? I’m doing a lot.’” G06, GP*

### 4. Clinical validation and reassurance

PCCs used A&G to validate their management of patients in primary care, which appeared to be necessary in the context of changing primary care landscape with increasing clinical complexity, and blurring of primary and secondary care task division.

> *“I think it’s [A&G use] probably increased. I think [I] probably use it more now as clinical situations are becoming more complicated, as waiting lists have increased massively. I think you, you know-to be able to contact a clinician, especially if they’re quite responsive [*…*] it is really quite reassuring, especially if you’ve done something with a patient, you know, in general practice since I’ve been a GP for 20 years, like the complexity of the patients that I’m seeing now that I’m expected to deal with, has changed vastly. You know, people are getting older. The comorbidities are increasing, the number of medications they’re on, you know, the different types of medications, the complications that go with those. G18, GP*

PCCs reported using A&G as a way to manage clinical risk during decision making, particularly for patients whose presentations did not exactly fit existing referral criteria.

> **“***If it’s something I’m not sure it needs a referral or say someone isn’t quite fitting criteria for referral, like a two week wait, but I’m worried there’s something underlying that we’re missing, it can be useful for that. G01, GP*

PCCs reported using A&G to provide clinical reassurance for decisions made in the boundaries between primary and secondary care remits.

> ***“****…I don’t mind dealing with this, but I just want to check that you think that’s appropriate and that I’m not sort of maybe outside of Primary Care’s remit here and it needs to be seen more in secondary care*.*’ G12, GP*

PCCs found this especially important where guidelines were potentially ambiguous about the extent of primary care remit. *“I find it very helpful. I think it provides me insurance, especially if you are doing things that are perhaps towards the top end or things not licenced for use that you can use them, type things. So it’s not your general NICE guidelines this is what you do. It tends to be the things on the fringe that you perhaps don’t know what you don’t know. or where you need confirmation that you can do things. G13*, GP

PCCs also used A&G as a tool to manage patient expectations, and to provide credibility and transparency to seeking support from specialists to confirm their management plans:

> *“I feel A&G gives it the credibility that the advice you’re getting is from a clinician who is in the specialty that you’re worried about, and secondly, it goes on the patients records*… *So I think yeah, definitely it brings a lot of confidence to your decision-making, provides confidence to the patient and provides credibility to what you’ve done. And it’s visible, it’s out there on the records. G04*, GP

## Discussion

### Summary

Our study, the first qualitative exploration of the views of PCCs on using the A&G system in the NHS, presents four overarching themes driving the decisions of PCC to use A&G for the care of their patients: clinical presentation of patients including acuity and complexity, the need to navigate through local healthcare systems, previous experience of A&G use and consequences on workload, and to validate their clinical decision-making. Barriers to successful integration of A&G into routine clinical practice included the need for appropriate turnaround times for specialist advice, and lack of clarity over the distribution of labour across the primary and secondary care interface, with the corresponding impact on primary care workload. Understanding use of A&G by primary care is critical for optimising the A&G process across the healthcare system.

#### Strengths and limitations

Our study has several strengths. Our large qualitative sample facilitated interviews with a range of professionals within the primary care team and included experienced and trainee clinicians allowing capture of views across the workforce. Recruiting from general practices across a spread of geographical regions enabled experiences from use of A&G with different NHS Trusts, and in practices across the socio-economic spectrum. Interviews were conducted by a non-clinical team which limited the possibility of personal influence from interviewers’ experience of using A&G.

During the study, the landscape around A&G changed; A&G was initially included in British Medical Association related collective action whereby the use of A&G could have been limited by individual practitioners,^29^ and later the UK Government announced that the use of A&G in primary care would be incentivised.^19^ This landscape is set to change further as A&G looks to be mandated in the England GP Contract 2026/27, though our common themes and key facilitators to optimise A&G use remain relevant.

#### Comparison with existing literature

Published literature on PCC decision making around the use of A&G is limited, however national guidance does exist in UK. Our qualitative findings suggest that PCCs decide to use A&G to obtain timely specialist input that clarifies or guides management and navigate sometimes complex local pathways. This aligns with national guidance that recommends A&G be used to obtain treatment advice, guide interpretation of results, and assess referral appropriateness, as well as to identify the most suitable service to use.^30^ Consistent with advice from the RCGP, PCCs value A&G when faced with diagnostic uncertainty and borderline referral thresholds. Contact with specialist colleagues to improve patient care and reduce burden, where safe, is appreciated. The place of A&G in shared decision making is less clear in our data.^31^

Internationally and in other healthcare systems, similar asynchronous advice seeking care processes similar to A&G have been developed and evaluated.^32-36^ For example, in Queensland Australia, Job et al have evaluated the use of ‘eConsultant.’ Qualitative interviews with 11 GPs and 4 stakeholders from the eConsultant service supported avoiding unnecessary referrals through timely access to specialist advice. Similar to our findings, barriers identified included competing priorities and the digital infrastructure and facilitators, relative advantage over other options in terms of patient benefit.^37^

Our data adds national nuance around the importance of prior experience in terms of timeliness and clinical helpfulness of responses as drivers of use. We also observed system constraints shaping decisions, reflective of research exploring collaboration between primary and secondary care professionals where gaps in continuity of care were highlighted as potential means of improving collaboration.^38^ A&G was observed to be used to reconcile local variability in healthcare systems, anticipated waits prompting the use of A&G to deliver interim care, and the possibility of an A&G request being converted into a referral effectively transformed its use to that of a triage tool (much like the referral advice system already established in the eRS). PCCs also reported times whereby A&G had been removed locally due to capacity limitations in secondary care to respond. Such implementation challenges may not be captured in the current national narrative and the voice of the Secondary Care Clinician (SCC) and indeed the patient has not yet been heard; these will be reported separately.

#### Implications for research and/or practice

Our study has found that PCCs do choose to use A&G to support patient care and clinician decision making about patient care. Addressing system processes including timeliness and clinical helpfulness of responses and enabling continued conversations within one care episode, and ensuring specialist responses include actions that are consistent with primary care remit will enable optimisation of a process that may be mandated as the exclusive route into secondary elective care. Understanding perspectives of SCCs, patients, and healthcare commissioners is needed to enable a cross-system approach to reducing compound pressures through A&G, whilst maintaining a safe and effective system that benefits patients and clinicians.

#### Transparency

The lead authors (AFN & RH) affirm that the manuscript is an honest, accurate, and transparent account of the study being reported; that no important aspects of the study have been omitted; and that any discrepancies from the study as originally planned (and, if relevant, registered) have been explained.

## Data Availability

The datasets generated during and/or analysed during the qualitative work packages will be available upon request from Alice Faux-Nightingale. Any subsequent requests for access to the data from anyone outside of the research team (e.g. collaboration, joint publication, data sharing requests from publishers) will follow the Keele University SOP data sharing procedure.

## Authors’ Contributions

All authors were involved in the writing, review & editing of the manuscript. Attributed author roles are summarised below.

*Conceptualisation and funding acquisition:* KJM, KPJ, RB, AFN, JH, RH, SLH, TH, CJ, CDM, VKW, CB

*Data curation, formal analysis and validation:* AFN, RH

*Methodology and project administration:* AFN, RH, CJ, CB, VW

*Supervision*: CJ, VKW, CB

## Acknowledgments

The authors would especially like to thank the members of the Patient Advisory Group members involved in BADGER, particularly John Haines as public co-applicant on the grant, for their valuable contributions to this study. We are also thankful to Keele PPI team for their support of the public contributors. We are grateful to Expert Citizens, Telford African & Afro-Caribbean Resource Centre and Open Harmony for their insights and support with study documents and interpretation of findings. The authors would like to thank the NIHR ARC West Midlands and the Study Oversight Committee for their support and the West Midlands and North West Regional Research Delivery Networks for their support with recruitment.

## Data sharing

The datasets generated during and/or analysed during the qualitative work packages will be available upon request from Alice Faux-Nightingale. Any subsequent requests for access to the data from anyone outside of the research team (e.g. collaboration, joint publication, data sharing requests from publishers) will follow the Keele University SOP data sharing procedure. Data Management and Access Plan

