## Supplementary material 2 for "Why Primary Care Clinicians use Advice and Guidance: A qualitative study"

**Building an evidence base for the use of ADvice and GuidancE Referrals at the primary-secondary care interface (BADGER), qualitative study - PCC Topic guide**

### 1. Background

- Please could you briefly outline your current role
  - How long have you been working in this role?
  - How long have you been using A&G for?
  - How often do you use A&G?
  - In your practice, who makes A&G referrals (GPs, ANPs, PAs and with what level supervision)?

### 2. Use of A&G

- What are your experiences of initiating and acting upon A&G requests?
  - In what kind of situations do you use A&G?
  - What are you hoping for when you send a request for A&G?
  - Do you offer patients a choice in their referral pathway?
  - Do you, and how do you do you explain A&G to patients?
  - Are there situations where you wouldn’t tell a patient you were using A&G in their care?
  - What do you include in a request for A&G?
  - What do you think you should include ideally?
  - How does using A&G influence the consultation and rapport patients?
- What does an ideal response to A&G look like?
  - How helpful are the responses that you receive?
    - How could they be improved?
- Are, and if so how, are the outcomes of A&G communicated back to patients?
- What is the process for using A&G in your practice?
  - How do you document and code A&G activity in the electronic healthcare record?
    - Has this process changed over time?
  - What measures are in place to ensure A&G is acted upon?
  - How do you perceive the role of the wider clinical team /additional role reimbursement staff
- Are there certain specialities or condition groups that you use A&G for more so than others?
  - If so, what are the reasons for this?
  - What specialities or condition groups do you tend not use A&G for?
- Are there patient groups who A&G is more suited for?
  - If so, what makes it more or less suitable for these patients?
- If the patient has multiple long term conditions (MLTC), severe mental illness (SMI), and / or learning disabilities (LD), how might this impact your decision about whether to use to use A&G?
  - Do MLTC, severe mental illness (SMI) and / or Learning Disabilities (LD) influence your decision to refer through particular pathways? (e.g. direct referral rather than A&G)
  - Does the presence of MLTC impact upon the A&G provided by SCSs?
- Could you describe an ‘ideal’ interaction with patients using A&G?

### 3. Perceptions of A&G

- Do you feel that A&G is clinically safe?
  - Are you able to share example of A&G has worked well – what were the measures of outcome (e.g. expedited pathway to diagnosis?)
  - Are you able to share examples of when A&G has not worked well – what were the measures of outcome (e.g. delayed diagnosis or unplanned admission)
- Do you perceive a transfer of clinical risk when you use A&G?
- One of the aims of A&G is to relieve pressure on the delivery of elective care – do you think this is the case?
- How does A&G use affect your workload?
  - When compared to making a routine referral, is the A&G process more or less burdensome on your time and activity?
- Does A&G use affect how you see your role?
  - Do you think it affects how patients see your role?
  - For example, does it mean working outside of your own competencies?
  - Do you feel A&G is a facilitator or barrier to patients accessing secondary care?
- How do patients perceive the use of A&G?
  - Do you explain to patients that you are going to use A&G in their care?
    - If so, in what ways do you phrase this?
    - If not, what are your reasons for this? Are there some patients who you would discuss A&G with? Why is this?
  - Do you think A&G is something patients should be actively aware of and involved in? What are your reasons for thinking this?
  - Do patients understand the remit of A&G?
    - e.g. do patients complain about not seeing a specialist? How do you manage patient expectations of the referral process?
